## supporting information for "A comparison of machine learning models versus clinical evaluation for mortality prediction in patients with sepsis"

**Affiliations**

**Supporting information**

**S1 supporting information. Extended description of clinical criteria and risk scores.**

**S2 supporting information. Background information on machine learning models reviewed in the current study.**

**Supporting Tables**

**S1 Table. Overview of variables present in the datasets.**

**S2 Table. Comparison of baseline statistical and machine learning models for predicting 31-day mortality risk.**

**S3 Table. Hyperparameters of XGBoost models.**

**S4 Table.** **Extended analysis of correlation between important model features and clinical risk scores.**

**S5 Table.** **Extended comparison of machine learning models with internal medicine physicians and clinical risk scores.**

**S6 Table. Inter-rater agreement of internal medicine physicians.**

**S7 Table. Machine learning comparison to alternating physician groups.**

**Supporting Figures**

**S1 Fig. Flow diagram of study inclusion.**

**S2 Fig. Five-fold cross validation of diagnostic performance of XGBoost models.**

**S3 Fig. Five-fold cross validation of calibration of XGBoost models.**

**S4 Fig. Receiver operating characteristic analysis of machine learning model, risk scores and internal medicine physicians.**

**S5 Fig. Individual performance of internal medicine physicians.**

**Supporting information**

**S1 supporting information. Extended description of clinical criteria and risk scores.**

In the current manuscript we describe several clinical criteria and risk scores. Below is a detailed description of each score or criteria and their application in our manuscript:

- **qSOFA:** the simple quick sequential organ failure assessment (qSOFA) is a bedside tool to identify patients with suspected infection who are at greater risk for a poor outcome. It uses three criteria, assigning one point for low blood pressure (systolic blood pressure ≤ 100 mmHg), high respiratory rate (≥22 breaths per min), or altered mentation (Glasgow coma scale < 15). In the current study we included patients with ≥ 2 points.
- **SIRS:** the systemic inflammatory response syndrome (SIRS) criteria has the same objective as the qSOFA criteria. SIRS assigns one point for tachycardia (heart rate > 90 beats/min), tachypnea (respiratory rate > 20 breaths/min), fever or hypothermia (temperature >38 or <36 °C), and leukocytosis, leukopenia, or bandemia (white blood cells > 12 * 10^9^/L , < 4 * 10^9^/L or bandemia ≥ 10%). In the current study we included patients with ≥2 points.
- **abbMEDS:** the abbreviated Mortality Emergency Department Sepsis (abbMEDS) score assesses sepsis severity and predicts mortality. This score assigns six points for terminal disease, three for respiratory difficulty (respiratory rate >30 breaths/min), three for septic shock, three for low thrombocytes (< 150 * 10^9^/L), three for a higher age (> 65 years), two for a lower respiratory tract infection, two for being a nursing home resident and two for an altered mental state (Glasgow coma scale < 15). abbMEDS is used in three categories: low (0 – 4 points), intermediate (5 – 12 points) and high risk (13 – 24), we used a cut-off value of 7 to dichotomize the outcome.
- **mREMS:** the modified rapid emergency medicine score (mREMS) is a triage score that is used to predict 28-day in-hospital mortality. This score assigns points for age (0: ≤44, 1: 45-64, 3: 65-74, 4: >74), systolic blood pressure (0: 110-159, 1: 160-199 and 90-109, 2: ≥200 and 80-89, 4: ≤79), heart rate (0: 70-109, 2: 110-139 and 55-69, 3: 140-179 and 40-54, 4: >179 and ≤39), respiratory rate (0: 12-24, 1: 25-34 and 10-11, 2: 6-9, 3: 35-49, 4: >49 and ≤5), oxygen saturation (0: ≥89, 1: 86-89, 3: 75-85, 4: <75) and the Glasgow coma scale (0: 14 or 15, 2: 8-13, 5: 5-7, 6: 3 or 4).
- **SOFA:** the SOFA-score was initially developed as a tool to learn from the evolution of organ failure in sepsis, but later was extensively validated to predict morbidity and mortality in several populations (Ceriani et al., 2003, Chest; Minne et al., 2008, Crit Care). It scores 1-4 points for each of the six organ systems, and we calculated it according to the formula described in the original paper (Vincent et al., 1996, Inte Care Med).

**S2 supporting information. Background information on machine learning models reviewed in the current study.**

We conducted a comparison of available algorithms on the 31-day mortality prediction task. We considered the following algorithms:

- **Logistic regression:** logistic regression is a statistical technique that in its basic form uses a logistic function to model a binary dependent variable. A simple logistic regression model was used with L2 regularization, a tolerance of 1^e-4^ and a maximum amount of iterations of 1,000. We used the limited Broyden–Fletcher–Goldfarb–Shanno (lbfgs) algorithm as optimizer. Logistic regression was implemented using Python (version 3.7.1) and the sklearn (version 0.22.1) package.
- **Multi-layer perceptron neural network:** neural networks are statistical models vaguely inspired by the biological neural networks that constitute animal brains. A simple feed-forward, multi-layer perceptron neural network was implemented consisting of three hidden layers with respectively 128, 64 and 32 neurons and ReLU activation functions. We trained the network using a constant learning rate of 0.001 with a batch size of 1 and the Adam optimization scheme. The neural network was implemented using Python programming language (version 3.7.1) using packages Keras (version 2.2.2) and scikit-learn (version 0.22.1).
- **Random Forest:** random forests is an ensemble learning method for classification that operate by constructing a multitude of decision trees at training time and outputting the class that is the mode of the classes (classification).A random forest classifier with a decision tree as base learner, consisting of 200 trees with gini criterion and a maximum depth of 50 was used. We used bootstrapped samples for building trees.
- **Gradient-boosting systems:** gradient boosting is a machine learning technique for classification problems which produce a prediction model in the form of an ensemble of weak prediction models, typically decision trees. In contrast to random forests, it builds the model in a stage-wise fashion like other boosting methods do**.** We used the XGBoost implementation of gradient-boosting systems. Each implementation has specific unique implementation details, but all use decision trees as the base weak learner and gradient boosting to iteratively fit a sequence of such trees. We used a learning rate of 0.075, a maximum number of trees of 300 and a maximum depth of each base learner to be 13. We implemented this using the Python programming language (version 3.7.1) using the package XGBoost (version 0.90).

**Supporting Tables**

**S1 Table. Overview of variables present in the datasets described in the manuscript.**

The laboratory dataset consisted exclusively of laboratory variables with age, sex and time of request. The laboratory and clinical dataset contained all variables from the laboratory dataset and additionally clinical and vital characteristics.

| **Laboratory dataset^1^** | | **Laboratory and clinical dataset** |
| --- | --- | --- |
| Age  Sex  Request time  Sodium bicarbonate  ALAT  Albumin  Alkaline phosphatase  Alpha-1-fetoprotein  Ammonia  Amylase  Anti Xa  Anti-thrombin  APTT  ASAT  Atypical lymphocytes  Base excess  Basophiles  Bilirubin  Blasts  Blood transfusion  Calcium ion  Calcium total  Chloride  CK  CKD-EPI  CK-MB  Cortisol  Creatinin  CRP  D-Dimers  Direct Antiglobulin Test  Dysmorphic erytrocytes  Eosinophil  Erythroblasts  Erythrocytes  Estradiol  Ferritin  Fibrinogen  Folic acid  Fragmentocytes  PSA  Free T4  Gamma GT  Gentamycin  Glucose  Haptoglobin  HbCO  HbO2 | HDL  Hematocrit  Hemoglobin  INR  Iron  Lactate  Lactate dehydrogenase  Leukocytes  Lipase  Lymphocytes  Magnesium  MCV  MDRD  Metamyelocyte  Monocyte  Myelocyte  Neutrophils  NT-proBNP  Osmolality  pCO2  pH  Phosphate  Platelet count  pO2  Poikilocytosis  Potassium  Promyelocytes  PT  PTH  Reticulocyte  Rod-like granulocytes  Sedimentation rate  Segment core granulocytes  Sodium  Specific gravity  Standard sodium bicarbonate  Total CO2  Total protein  Toxic grain  Transferrin  Transferrin saturation  Triglycerides  Troponin T  TSH  Urea  Uric acid  Urobilinogen  Vitamin 25 (OH) D3  Vitamin B12 | Laboratory dataset variables  Weight  Length  Policy restrictions  Respiratory rate  Saturation  Temperature  Glasgow coma score  FIO2  Heart rate  Systolic BP  Diastolic BP  Inotropics/vasopressors  ECG rhythm |

^1^ For each of the laboratory variables we generated an additional binary ‘absence’ or ‘presence’ variable representing whether or not this laboratory parameter was requested.

**S2 Table. Comparison of baseline statistical and machine learning models for predicting 31-day mortality risk.**

We performed a baseline comparison of statistical and machine learning models (S1 supporting information) for the 31-day mortality prediction task using the laboratory dataset. We used five-fold cross validation to assess model performance. Performance was assessed by area under the receiver operating characteristic curve (AUC) and accuracy.

| **Evaluation**  **metric** | **Logistic regression** | **Multi-layer perceptron** | **Random Forest** | **XGBoost** |
| --- | --- | --- | --- | --- |
| AUC | 0.633  (0.606 – 0.660) | 0.658  (0.632 – 0.685) | 0.723  (0.689 – 0.756) | **0.813**  **(0.791 – 0.835)** |
| Accuracy | 0.826  (0.820 – 0.833) | 0.868  (0.858 – 0.877) | 0.842  (0.831 – 0.853) | **0.873**  **(0.864 – 0.883)** |

**S3 Table. Hyperparameters of XGBoost models.**

Hyperparameters were based on theoretical reasoning rather than hyperparameter tuning. This was done to prevent overfitting on hyperparameters due to small sample size. “Base_score”, “Missing”, “Reg_alpha”, “Reg_lambda” and “Subsample” parameters were standard values provided by the XGBoost interface. “Max_depth”, “max_delta_step” and “estimators” were values we internally use for these kind of machine learning models. During the study, hyperparameters were never adjusted to gain performance in our validation dataset.

| **Hyperparameter** | **Value** | **Explanation** |
| --- | --- | --- |
| Max_depth | 13 | Determines how deeply each tree is allowed to grow during any boosting round. |
| Max_delta_step | 3 | Maximum delta step we allow each tree’s weight estimation to be. |
| Learning rate | 0.075 | Degree to which weights are adjusted each learning iteration. |
| Base_score | 0.5 | Initial prediction score of all instances (global bias). |
| Missing | N/A | Value which is represented as missing. Put onto N/A as no imputation was performed in data processing. |
| Reg_alpha | 0 | L1 regularization term on weights |
| Reg_lambda | 1 | L2 regularization term on weights |
| Subsample | 1 | Percentage of samples used per tree; low value can lead to underfitting. |
| Estimators | 300 | Number of trees you want to build. |

**S4 Table.** **Extended analysis of correlation between important model features and clinical risk scores.**

To study the correlation between the most important features contributing to model predictions and the clinical criteria (qSOFA and SIRS) and risk scores (abbMEDS and mREMS), we compared their existence in both. The top-20 most important features (Fig. 3 in manuscript) are compared to all criteria in the clinical scores (S1 supporting information). Most of the features present in the clinical criteria and scores are also among the most important features in the lab + clinical machine learning model.

| **Lab**  **model top-20 features** | **Clinical criteria and scores** | | | | **Lab/ clinical**  **model top-20 features** | **Clinical criteria and scores** | | | |
| --- | --- | --- | --- | --- | --- | --- | --- | --- | --- |
|  | SIRS  0/4 | qSOFA  0/3 | MEDS  2/6 | REMS  1/6 |  | SIRS  3/4 | qSOFA  3/3 | MEDS  4/6 | REMS  5/6 |
| 1. Blood group (ordered) |  |  |  |  | 1. Heart rate | X |  |  | X |
| 2. Urea |  |  |  |  | 2. Blood group (ordered) |  |  |  |  |
| 3. Albumin |  |  |  |  | 3. Urea |  |  |  |  |
| 4. CKD-EPI |  |  |  |  | 4. Albumin |  |  |  |  |
| 5. Blood group |  |  |  |  | 5. Magnesium |  |  |  |  |
| 6. Platelet count |  |  | X |  | 6. Platelet count |  |  | X |  |
| 7. Age |  |  | X | X | 7. GCS |  | X | X | X |
| 8. Total protein |  |  |  |  | 8. Oxygen saturation | X | X | X | X |
| 9. CRP |  |  |  |  | 9. Age |  |  | X | X |
| 10. Sodium |  |  |  |  | 10. Temperature | X |  |  |  |
| 11. Glucose (arterial) |  |  |  |  | 11. Glucose |  |  |  |  |
| 12. LD |  |  |  |  | 12. Systolic BP |  | X |  | X |
| 13. Lactacte |  |  |  |  | 13. Creatinine |  |  |  |  |
| 14. Creatinine |  |  |  |  | 14. Amount of lab |  |  |  |  |
| 15. Lipase |  |  |  |  | 15. CRP |  |  |  |  |
| 16. Bilirubin |  |  |  |  | 16. ALAT |  |  |  |  |
| 17. Gamma-GT |  |  |  |  | 17. A. Fib (history) |  |  |  |  |
| 18. Alk. phosphatase |  |  |  |  | 18. CO2 (arterial) |  |  |  |  |
| 19. Magnesium |  |  |  |  | 19. Bilirubin |  |  |  |  |
| 20. Hemoglobin |  |  |  |  | 20. Calcium |  |  |  |  |

**S5 Table.** **Extended comparison of machine learning model with internal medicine physicians and clinical risk scores.**

In addition to sensitivity and specificity, we evaluated the performance of each group by positive predictive value (PPV), negative predictive value (NPV), accuracy and area-under-the receiver operating characteristics curve (AUC). The machine learning model shows superior performance in each of these metrics, which is consistent with the findings presented in the manuscript. Confidence intervals were calculated using binomial testing and AUC’s were compared using DeLong’s test.

| **Evaluation**  **metric** | **XGBoost**  **model** | **abbMEDS** | **mREMS** | **SOFA** | **Internal medicine physicians** |
| --- | --- | --- | --- | --- | --- |
| Sensitivity, % | 92.3  (87.1 – 95.3) | 53.8  (44.1 – 63.6) | 61.5  (52.0 – 71.1) | 76.9  (68.7 – 85.2) | 72.1  (61.3 – 82.2) |
| Specificity, % | 78.2  (70.1 – 86.3) | 72.4  (63.7 – 81.2) | 64.4  (55.0 – 73.8) | 73.6  (64.9 – 82.2) | 74.2  (63.9 – 82.1) |
| PPV, % | 38.7  (29.2 – 48.3) | 22.6  (14.4 – 30.8) | 20.5  (12.6 – 28.4) | 30.3  (21.3 – 39.3) | 29.5  (20.5 – 38.4) |
| NPV, % | 98.6  (96.2 – 100.0) | 91.3  (85.8 – 96.8) | 91.8  (86.4 – 97.2) | 95.5  (91.5 – 99.6) | 94.8  (90.5 – 99.2) |
| Accuracy | 0.800  (0.722 – 0.878) | 0.700  (0.610 – 0.790) | 0.640  (0.546 – 0.734) | 0.740  (0.654 – 0.826) | 0.738  (0.651 – 0.824) |
| AUC | 0.852  (0.783 – 0.922) | 0.631  (0.537 – 0.726) | 0.630  (0.535 – 0.724) | 0.752  (0.667 – 0.836) | 0.735  (0.648 – 0.821) |
| P-value | N/A | 0.021 | 0.016 | 0.042 | 0.189; 0.072; 0.068; 0.032^a^ |

*^a^ Individual P-values were calculated for each of the internal medicine physicians.*

**S6 Table. Inter-rater agreement of internal medicine physicians.**

Cohen’s kappa was used to measure the inter-rater agreement between the internal medicine physicians. The level of agreement was interpreted as nil if κ was 0 to 0.20; minimal, 0.21 to 0.39; weak, 0.40 to 0.59; moderate, 0.60 to 0.79; strong, 0.80 to 0.90; and almost perfect, 0.90 to 1.0.

| **Internist** | 1 (consultant) | 2 (consultant) | 3 (resident) | 4 (resident) |
| --- | --- | --- | --- | --- |
| 1 (consultant) | - | 0.52 | 0.54 | 0.46 |
| 2 (consultant) | 0.52 | - | 0.61 | 0.67 |
| 3 (resident) | 0.54 | 0.61 | - | 0.63 |
| 4 (resident) | 0.46 | 0.67 | 0.63 | - |

**S7 Table. Machine learning comparison to alternating physician groups.**

In each comparison between the machine learning model and the physicians group, a single physician was removed from the physician group. In every comparison the machine learning model outperforms the physicians. This analysis shows that the higher performance of the machine learning model was not due to systemic underperformance of a single physician.

| **Group** | **Sensitivity** | | **Specificity** | |
| --- | --- | --- | --- | --- |
|  | Mean (95% CI) | *P-value* compared to model | Mean (95% CI) | *P-value* compared to model |
| All internists | 0.72  [0.62-0.81] | <0.001 | 0.74  [0.64-0.82] | 0.509 |
| Internist 2, 3, 4 | 0.71  [0.61-0.80] | <0.001 | 0.75  [0.65-0.83] | 0.524 |
| Internist 1, 3, 4 | 0.69  [0.60-0.78] | <0.001 | 0.76  [0.67-0.84] | 0.653 |
| Internist 1, 2, 4 | 0.74  [0.66-0.83] | 0.001 | 0.74  [0.65-0.82] | 0.447 |
| Internist 1, 2, 3 | 0.77  [0.69 – 0.85] | 0.003 | 0.72  [0.63-0.81] | 0.316 |

**Supporting Figures**

**S1 Fig. Flow diagram of study inclusion.**

During the study period 5,967 patients that presented to our emergency department were referred to an internal medicine physician. Of these patients, 1420 patients fulfilled two or more SIRS and/or qSOFA criteria. After exclusion of 76 patients, a number of 1,344 patients were separated into development and validation datasets.

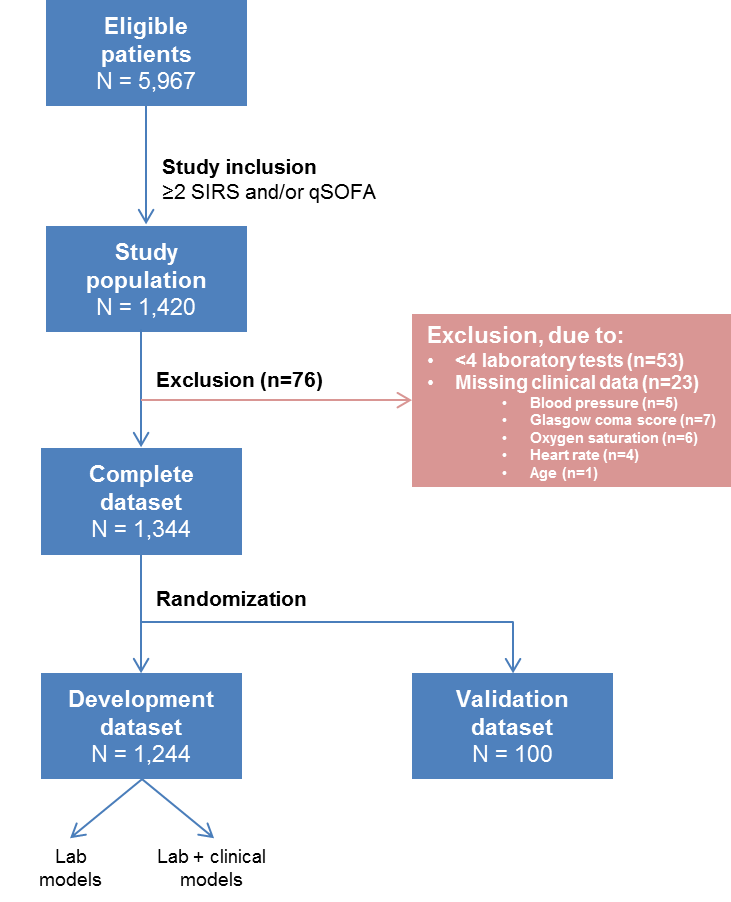

**S2 Fig. Five-fold cross validation of diagnostic performance of XGBoost models.**

During each fold of cross-validation, we assessed predictive performance by area under the receiver operating characteristic curves (AUC). Performance was determined for models trained with laboratory data (A) and models trained with laboratory + clinical data (B) to predict 31-day mortality.

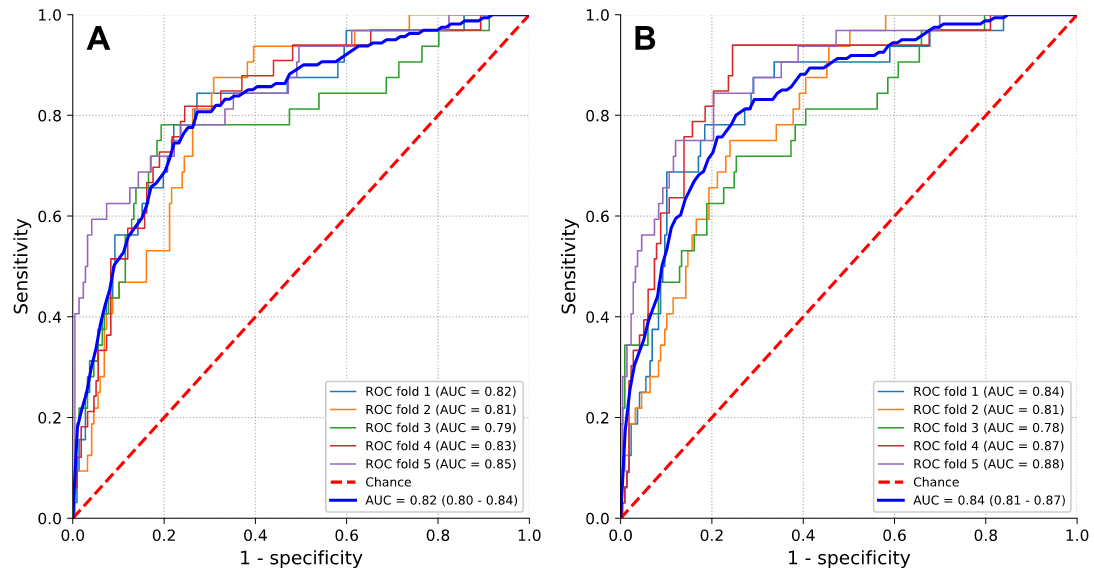

**S3 Fig. Five-fold cross validation of calibration of XGBoost models.**

During each fold of cross-validation, we assessed calibration by calibration curves and their respective brier scores. Calibration was determined for models trained with laboratory data (A) and models trained with laboratory + clinical data (B).

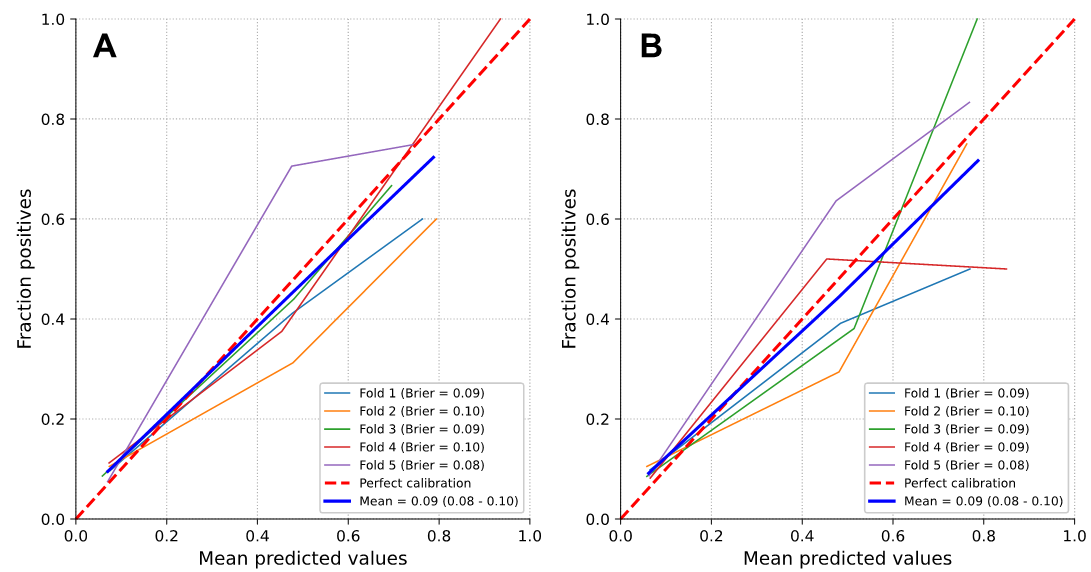

**S4 Fig. Receiver operating characteristic analysis of machine learning model, risk scores and internal medicine physicians.**

Receiver operating characteristics analysis of the lab + clinical machine learning model (AUC: 0.852 [0.783-0.922]), abbMEDS (0.631 [0.537-0.726]), mREMS (0.630 [0.535-0.724]), SOFA (AUC: 0.752 [0.667 – 0.836]) and internal medicine physicians (mean 0.735 [0.648-0.821]). Internal medicine physicians were depicted as bullets in the ROC analysis.

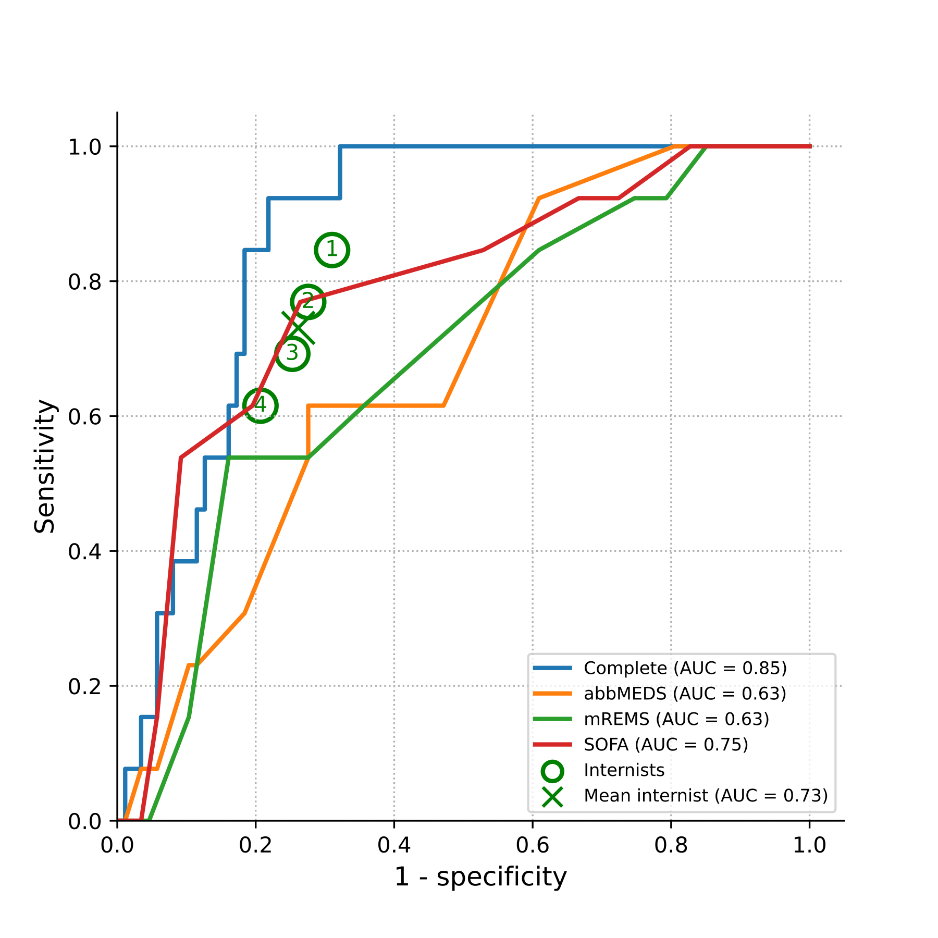

**S5 Fig. Individual performance of internal medicine physicians.**

Predictive performance of all internal medicine specialists (n=4; 2 experienced consultants in acute internal medicine and 2 experienced residents acute internal medicine) was assessed by sensitivity (left) and specificity (right). Consultants are depicted in grey and residents in orange.

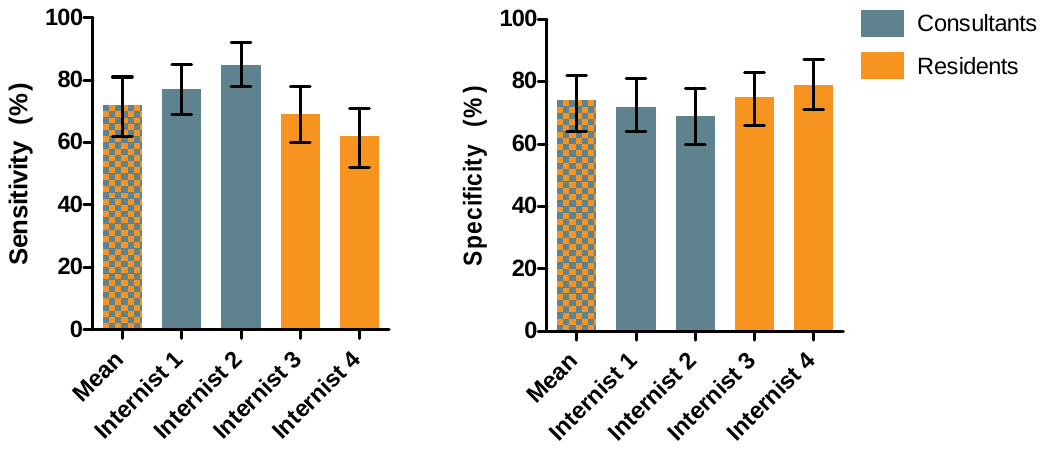
